# SARS-CoV-2 and influenza co-infection throughout the COVID-19 pandemic: An assessment of co-infection rates and cohort characterization

**DOI:** 10.1101/2022.02.02.22270324

**Authors:** Colin Pawlowski, Eli Silvert, John C. O’Horo, Patrick J. Lenehan, Doug Challener, Esteban Gnass, Karthik Murugadoss, Jason Ross, Leigh Speicher, Holly Geyer, AJ Venkatakrishnan, Andrew Badley, Venky Soundararajan

**Author notes:** Correspondence: Venky Soundararajan & Andrew Badley. These authors contributed equally (Colin Pawlowski, Eli Silvert, John C. O’Horo).

## Abstract

**Background:** Case reports of patients infected with COVID-19 and influenza virus (“flurona”) have raised questions around the prevalence and clinical significance of these reports.

**Methods:** Epidemiological data from the *HHS Protect Public Data Hub* was analyzed to show trends in SARS-CoV-2 and influenza co-infection-related hospitalizations in the United States in relation to SARS-CoV-2 and influenza strain data from *NCBI Virus* and *FluView*. In addition, we retrospectively analyzed all cases of PCR-confirmed SARS-CoV-2 across the *Mayo Clinic Enterprise* from January 2020 to January 2022 and identified cases of influenza co-infections within two weeks of PCR-positive diagnosis date. Using a cohort from the Mayo Clinic with joint PCR testing data, we estimated the expected number of co-infection cases given the background prevalences of COVID-19 and influenza during the Wuhan (Original), Alpha, Delta, and Omicron waves of the pandemic.

**Findings:** Considering data from all states of the United States using HHS Protect Public Data Hub, hospitalizations due to influenza co-infection with SARS-CoV-2 were seen to be highest in January 2022 compared to all previous months during the COVID-19 pandemic. Among 171,639 SARS-CoV-2-positive cases analyzed at Mayo Clinic between January 2020 and January 2022, only 73 cases of influenza co-infection were observed. Identified coinfected patients were relatively young (mean age: 28.4 years), predominantly male, and had few comorbidities. During the Delta era (June 16, 2021 to December 13, 2021), there were 9 lab-confirmed co-infection cases observed compared to 13.9 expected cases (95% CI: [12.7, 15.2]), and during the Omicron era (December 14, 2021 to January 17, 2022), there were 54 lab-confirmed co-infection cases compared to 80.9 expected cases (95% CI: [76.6, 85.1]).

**Conclusions:** Reported co-infections of SARS-CoV-2 and influenza are rare. These co-infections have occurred throughout the COVID-19 pandemic and their prevalence can be explained by background rates of COVID-19 and influenza infection. Preliminary assessment of longitudinal EHR data suggests that most co-infections so far have been observed among relatively young and healthy patients. Further analysis is needed to assess the outcomes of “flurona” among subpopulations with risk factors for severe COVID-19 such as older age, obesity, and immunocompromised status.

**Significance Statement:** Reports of COVID-19 and influenza co-infections (“flurona”) have raised concern in recent months as both COVID-19 and influenza cases have increased to significant levels in the US. Here, we analyze trends in co-infection cases over the course of the pandemic to show that these co-infection cases are expected given the background prevalences of COVID-19 and influenza independently. In addition, from an initial analysis of these co-infection cases which have been observed at the Mayo Clinic, we find that these co-infection cases are extremely rare and have mostly been observed in relatively young, healthy patients.

## Introduction

Since the beginning of the COVID-19 pandemic there have been lingering concerns around the possibility of a “twindemic” with influenza^1^, particularly as the COVID-19 pandemic extends through influenza seasons. Amidst the ongoing surge of Omicron-associated COVID-19 cases, recent reports of patients testing positive for both COVID-19 and influenza, dubbed as “flurona” patients^2^, have raised an alarm. Co-infection with SARS-CoV-2 and Influenza has been reported since early in the pandemic. A meta-analysis of co-infection prevalence studies from December 2019 to September 2020 found 79 individuals with concurrent COVID-19 and influenza infection among a total of 3,070 COVID-19 cases^3^. Some studies have reported that up to 20% of COVID-19 cases can demonstrate co-infection with other respiratory viruses.^4^ However, there is a lack of understanding of the significance of COVID-19 and influenza co-infections.

Epidemiological data on SARS-CoV-2, influenza, and co-infection-related hospitalizations in the United States is available from the HHS Protect Public Data Hub^5^. The HHS Protect Public Data Hub is a central COVID-19 data repository which aggregates United States healthcare data from various Health and Human Services (HHS) operating divisions including: Centers for Disease Control and Prevention (CDC), Centers for Medicare and Medicaid Services (CMS), Health Resources and Services Division (HRSD), and others. Availability of this national-level data allows us to investigate the temporal and geographic trends associated with COVID-19 and Influenza co-infections. Other data repositories such as NCBI Virus^6^ and FluView^7^ provide valuable data on SARS-CoV-2 and influenza strain prevalence in the United States. In addition, understanding the clinical characteristics and outcomes associated with COVID-19 and influenza co-infections requires a longitudinal analysis of data from sources such as clinical trials or electronic health records (EHRs) which have patient-level information. Previously, analyses of EHR data from the Mayo Clinic Enterprise have been used to assess various characteristics of COVID-19 symptomatology^8^, duration of infection^9^, and COVID-associated complications^10,11^.

In this study, we analyze epidemiological data from HHS Protect Public Data Hub in order to evaluate trends in COVID-19, influenza, and co-infection cases across the entire United States in relation to trends in SARS-CoV-2 and influenza strain prevalences. In addition, we apply machine-augmented curation models (neural networks) and other techniques to conduct an observational study over the Mayo Clinic Enterprise EHR database to assess the prevalence and clinical characteristics of co-infections from SARS-CoV-2 and influenza viruses in this multi-state health system.

## Methods

### Tracking temporal and geographic trends in SARS-CoV-2 and influenza co-infection-related hospitalizations in the United States

In order to track temporal trends in SARS-CoV-2, influenza, and co-infection-related hospitalizations in the United States over the course of the pandemic, we used the *“COVID-19 Reported Patient Impact and Hospital Capacity by Facility*” dataset^12^. The hospital population includes all hospitals registered with Centers for Medicare & Medicaid Services (CMS) as of June 1, 2020. For each reporting hospital, this dataset includes 7-day average case counts of hospitalized patients with PCR-confirmed COVID-19, PCR-confirmed influenza, and co-infections. We considered a subset of these hospitals with data available for these variables each week from October 30, 2020 to January 13, 2022. In addition, we filtered out hospitals with inconsistent data (e.g. more co-infection hospitalized cases than total influenza hospitalized cases for a single week) to obtain a final set of 3,003 hospitals for analysis. Since case counts less than 4 individuals are censored in this dataset, we imputed these values to be 1 for the computation of averages. For each week, we summed the average case counts across all 3,003 hospitals to obtain estimates for the average number of hospitalized patients with SARS-CoV-2, influenza, and co-infections at these facilities during that week.

To track geographic trends in SARS-CoV-2, influenza, and co-infection-related hospitalizations in the United States over the course of the pandemic, we used the *“COVID-19 Reported Patient Impact and Hospital Capacity by State Timeseries”* dataset^13^. This is similar to the above dataset except the data is aggregated at the state level, and case counts for facilities with fewer than 4 individuals are not censored for the sums. For each state on a map, we show the total number of patients hospitalized with laboratory-confirmed COVID-19 and influenza co-infections across all of the reporting facilities on January 15, 2022. In addition, for each state we compute the percentage of COVID-19 and influenza co-infections as the total number of hospitalized patients with co-infections divided by the total number of patients hospitalized with COVID-19.

### Mayo Clinic Institutional Review Board

We performed a retrospective analysis on electronic health record (EHR) data from the Mayo Clinic under the IRB #20-003278 *“Study of COVID-19 patient characteristics with augmented curation of Electronic Health Records (EHR) to inform strategic and operational decisions*.*”* This includes all individuals with research authorization on file who received a PCR test for SARS-CoV-2 at Mayo Clinic sites since January 1, 2020. The Mayo Clinic Enterprise is a multi-state academic medical center with major campuses in Rochester, MN, Jacksonville, FL, and Scottsdale, AZ, along with additional satellite sites in other states including Iowa and Wisconsin.

### Cohort definitions

The study population included all individuals who have received a positive PCR test for SARS-CoV-2 at the Mayo Clinic between January 1, 2020 and January 17, 2022. From this study population, we constructed the following cohorts: (1) “Overall COVID-19”: all individuals with a positive PCR test for SARS-CoV-2, (2) “COVID-19 + Flu”: all individuals with a positive PCR test for SARS-CoV-2 along with a diagnosis of influenza within 14 days.

By definition, all individuals in the study population were included in the “Overall COVID-19” cohort. In addition, some individuals in the study population were included multiple times in this cohort if they had multiple positive PCR tests for SARS-CoV-2 during the study period spaced at least 90 days apart. For each individual, the date of the first positive PCR test was considered to be the primary infection, and the date of each subsequent positive PCR test spaced at least 90 days apart was considered to be a re-infection.

To construct the “COVID-19 + Flu” cohort, we used a combination of data sources from within Mayo Clinic Enterprise to determine influenza diagnosis: laboratory tests (**Table S1**), diagnostic billing codes (**Table S2**), and unstructured clinical notes. Cases without a laboratory correlate were manually reviewed to confirm influenza diagnosis. If any of these data sources indicated that an individual in the “Overall COVID-19” cohort had an influenza diagnosis within +/-14 days of their PCR test date, then this individual was included in the “COVID-19 + Flu” cohort. If there was conflicting information across the data types, i.e. there was a negative influenza lab test result within +/-7 days of a diagnostic billing code or diagnosis based on the notes, this was not considered to be a case of co-infection. Similar to the “Overall COVID-19” cohort, individuals may be included multiple times in the “COVID-19 + Flu” cohort if they had multiple co-occurrences of SARS-CoV-2 and influenza infection. The methodology to determine influenza diagnoses from the unstructured clinical notes is described below.

### Augmented curation to determine influenza diagnosis from clinical notes

We used a BERT-based neural network model^14^ to extract the sentiment of all mentions of influenza in the clinical notes for the study population. This phenotype sentiment model has been previously applied to determine signs and symptoms of COVID-19 from unstructured clinical notes with an out-of-sample accuracy of 93.6% and precision/recall values above 95%^8^. For each occurrence of a phenotype in a clinical note, the model outputs one of the following labels: “Yes”: confirmed diagnosis of the phenotype, “Maybe”: uncertain/differential diagnosis of the phenotype, “No”: ruled-out diagnosis of the phenotype, or “Other”: all other mentions of the phenotype (e.g. family history). For this application, we considered the following terms as synonyms for influenza: “influenza”, “influenza A”, “influenza B”, “influenza A/B”, “influenza virus”, “flu”, “avian flu”, “H1N1”, “H5N1”, “H3N2”, and “H7N9”. Individuals with at least one clinical note labeled “Yes” by the model with ≥ 90% confidence within +/-14 days of their positive PCR test were counted as positive for influenza and included in the “COVID-19 + Flu” cohort. Cases without laboratory correlates were manually reviewed and confirmed for inclusion.

### Curation of clinical covariates from structured EHR data

For each cohort, we curated clinical covariates from the structured EHR data with features including: demographics (age, sex, race, ethnicity), geographic location and time of PCR test, COVID-19 vaccination status, influenza vaccination status, and comorbidities, and 30-day clinical outcomes including: hospitalization, ICU admission, and mortality. For each individual case in each cohort, we considered the date of the positive PCR test for SARS-CoV-2 to be the index date (day = 0) to define the clinical covariates. To classify the geographic location of PCR testing, we used the following Mayo Clinic regions: Mayo Clinic - Arizona (includes PCR tests administered at the major campus in Scottsdale, AZ), Mayo Clinic - Florida (includes PCR tests administered at the major campus in Jacksonville, FL), and Mayo Clinic - Midwest (includes PCR tests administered at the major campus in Rochester, MN as well as surrounding Mayo Clinic Health Systems sites in Minnesota, Wisconsin, and Iowa).

To determine COVID-19 vaccination status, we considered the following FDA-authorized vaccines: Janssen (Ad26.COV2.S), Moderna (mRNA-1273), and Pfizer/BioNTech (BNT162b2). The categories for COVID-19 vaccination status were defined as: “Unvaccinated”: individuals who do not have an FDA-authorized COVID-19 vaccine on record, “Partial”: individuals who have received exactly 1 dose of either BNT162b2 or mRNA-1273, “Full”: individuals who have received exactly 1 dose of Ad26.COV2.S or exactly 2 doses of either BNT162b2 or mRNA-1273 at least 14 days prior to their PCR date, and “Boosted”: individuals who are fully vaccinated and have received an additional dose of either Ad26.COV2.S, BNT162b2, or mRNA-1273 at least 14 days prior to their PCR date. For each individual case, vaccination status is determined at the time of the associated positive PCR test, so it is possible that some individuals who are fully vaccinated now may be considered unvaccinated on their index dates. To determine influenza vaccination status, we checked if the individual received an influenza vaccine at least 14 days prior to their positive PCR test for SARS-CoV-2 within the current flu season beginning on August 1st. For example, an individual with a positive PCR test on October 15, 2021 would need to have received a flu vaccine between August 1, 2021 and October 1, 2021 in order to be considered vaccinated for the flu season. In **Table S3**, we provide a comprehensive list of the influenza vaccines considered.

We considered the following time periods of PCR testing dates: March 12, 2020 to March 15, 2021; March 16, 2021 to June 15, 2021; June 16, 2021 to December 13, 2021; and December 14, 2021 onwards. These time periods roughly correspond to the time periods when the SARS-CoV-2 strains Wuhan (original), Alpha, Delta, and Omicron were the dominant strains in the United States over the course of the study.

To determine the comorbidities for each cohort, we considered all 31 disease categories in the Elixhauser comorbidity index^15^, including: congestive heart failure, cardiac arrhythmia, valvular disease, pulmonary circulation disorder, peripheral vascular disorder, hypertension (uncomplicated), hypertension (complicated), paralysis, other neurological disorder, chronic pulmonary disease, diabetes (uncomplicated), diabetes (complicated), hypothyroidism, renal failure, liver disease, peptic ulcer disease (excluding bleeding), AIDS / HIV, lymphoma, metastatic cancer, solid tumor without metastasis, rheumatoid arthritis, coagulopathy, obesity, weight loss, fluid and electrolyte disorders, blood loss anemia, deficiency anemia, alcohol abuse, drug abuse, psychoses, and depression. For each disease category, individuals with an associated ICD-10 code within the past 5 years prior to their positive PCR testing date were counted as positive for the phenotype. For each individual, a single Elixhauser Comorbidity Index score was calculated using the Van Walraven method^16^.

### SARS-CoV-2 variant prevalence from the NCBI Virus database

We used the NCBI Virus database^6^ to determine the prevalence of the different SARS-CoV-2 variants over time. This dataset allows us to find the times of dominance for each variant and evaluate the most likely SARS-CoV-2 variant contributing to co-infection cases.

From the NCBI Virus database, we retrieved 3,483,717 SARS-CoV-2 genome samples from human hosts in the United States which were collected from January 1, 2020 to January 15, 2022 and span 1,385 PANGO lineages. We categorized the samples as an Alpha, Beta, Delta, Gamma, or Omicron variant using the variant classification information from the CDC to map the PANGO lineages to particular variants.^17^ We found the prevalence of the variants on each day in our study period by normalizing the daily variant sample count by the daily total sample count in the NCBI Virus dataset. In addition, we used this dataset to determine the dates at which a new variant became the dominant strain in the United States (i.e. the first date that Alpha was more prevalent than the ancestral lineage and the first date that Delta was more prevalent than Alpha).

### Influenza strain prevalence from the CDC FluView database

Similarly, we used the CDC FluView database^7^ to determine the prevalences of the two main influenza strains (A(H1N1)pdm09 and A(H3N2)) over time. This database provides the number of influenza strain types of all those sampled from sites in the United States on a weekly basis. From this database, we retrieved 38,052 influenza samples with known subtyping from January 1, 2020 to January 15, 2022. The 6 strains considered were A(H3N2v), A(H1N1)pdm09, A(H3N2), B, BVIC, BYAM. We computed the prevalence of the strains for each week in our study period as the weekly strain sample count divided by the weekly total subtype sample count.

### Estimates of COVID-19 and influenza co-infection prevalences

To estimate the prevalence of reported COVID-19 and influenza co-infections, we divided the number of reported COVID-19 and influenza cases by the total number of COVID-19 cases observed in the Mayo Clinic EHR database. In addition, to estimate the total (reported + unreported) prevalence of COVID-19 and influenza co-infections, we divided the number of observed COVID-19 and influenza co-infections (from lab tests only) by the number of COVID-19 cases with lab tests for influenza available. Finally, to estimate the prevalence of COVID-19 co-infections among all influenza cases, we divided the number of reported COVID-19 and influenza cases by the total number of influenza cases observed during the study period. For each estimate, 95% confidence intervals using Wilson’s score method^18^ are reported, which were computed using the “stats” package (version 4.1.2) in R.

### Estimates of the expected number of co-infection cases

For each of the study time periods, we estimated the expected number of COVID-19 and influenza co-infection cases based upon the background incidence rates of COVID-19 and influenza at the Mayo Clinic. We considered all cases at the Mayo Clinic during each study time period with PCR testing data available for both COVID-19 and influenza within +/- 14 days, including both positive and negative PCR tests. We considered the same time periods as described above: March 12, 2020 to March 15, 2021; March 16, 2021 to June 15, 2021; June 16, 2021 to December 13, 2021; and December 14, 2021 onwards. For each case, the date of the SARS-CoV-2 PCR test was used to determine the time period.

For each time period, we estimated the probability of COVID-19 among the co-tested population as the number of co-tested cases with positive PCR tests for SARS-CoV-2 divided by the total number of co-tested cases. Similarly, we estimated the probability of Influenza among the co-tested population as the number of co-tested cases with positive PCR tests for Influenza divided by the total number of co-tested cases. We compute the expected number of COVID-19 and Influenza cases for the time period as follows:

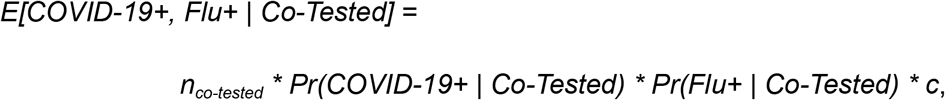

where:

*n*_*co-tested*_: Total number of co-tested cases during the time period,

*Pr(COVID-19*+ | *Co-Tested)*: Probability of COVID-19 among co-tested population for the time period,

*Pr(Flu*+ | *Co-Tested)*: Probability of influenza among the co-tested population for the time period, *c*: Estimated model parameter.

In the above estimate, the main assumption is that the probabilities of testing positive for COVID-19 and influenza are independent in the general population. In addition, we assume that the estimated model parameter *c* is constant over the course of the study. For these estimates of expected co-infection cases, we compute 95% confidence intervals using the Delta method^19^. In the following section, we provide a detailed description of the probability model underlying this formula along with a description of the method used to estimate the model parameter *c*.

### Probability model used to estimate the expected number of co-infection cases

In this section, we provide a description of the probability model which is used to estimate the expected number of COVID-19 and influenza co-infection cases at the Mayo Clinic. The primary assumption of this model is that the probabilities of testing positive for COVID-19 and influenza are independent in the general population of patients at the Mayo Clinic. For example, if the background prevalences of COVID-19 and influenza in the general population were 1% and 0.2% respectively, then we would expect the prevalence of co-infections to be 1% * 0.2% = 0.002%.

For each individual case, we define the index date to be the date of the SARS-CoV-2 PCR test. We define the following probability events:

*COVID-19*+: The individual received a positive PCR test for COVID-19 within +/- 14 days of the index date,

*Flu*+: The individual received a positive PCR test for influenza within +/- 14 days of the index date, and

*Co-Tested:* The individual received PCR tests for both COVID-19 and influenza within +/- 14 days of the index date.

Given these events, we define the following probabilities of COVID-19 and influenza mono-infections:

*Pr(COVID-19*+): Probability that the individual received a positive PCR test for COVID-19 within +/- 14 days of the index date (among patients with and without PCR testing),

*Pr(COVID-19*+ | *Flu*+): Probability that the individual received a positive PCR test for COVID-19 within +/- 14 days of the index date (among patients with positive PCR tests for influenza),

*Pr(COVID-19*+ | *Co-Tested)*: Probability that the individual received a positive PCR test for COVID-19 within +/- 14 days of the index date (among patients with PCR tests available for both influenza and COVID-19),

*Pr(COVID-19*+ | *Flu*+, *Co-Tested)*: Probability that the individual received a positive PCR test for COVID-19 within +/- 14 days of the index date (among patients with a positive PCR test result for influenza and a COVID-19 PCR test result available).

*Pr(Flu*+): Probability that the individual received a positive PCR test for influenza within +/- 14 days of the index date (among patients with and without PCR testing),

*Pr(Flu*+ | *COVID-19*+): Probability that the individual received a positive PCR test for influenza within +/- 14 days of the index date (among patients with positive PCR tests for COVID-19),

*Pr(Flu*+ | *Co-Tested)*: Probability that the individual received a positive PCR test for influenza within +/- 14 days of the index date (among patients with PCR tests available for both influenza and COVID-19).

*Pr(Flu*+ | *COVID-19*+, *Co-Tested)*: Probability that the individual received a positive PCR test for influenza within +/- 14 days of the index date (among patients with a positive PCR test result for COVID-19 and an influenza PCR test result available).

From the independence assumption, it follows that:

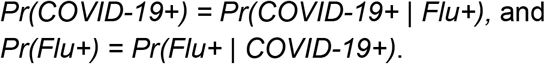

In addition, we define the following probabilities of COVID-19 and influenza co-infections:

*Pr(COVID-19*+, *Flu*+): Probability that the individual received positive PCR tests for both COVID-19 and influenza within +/- 14 days of the index date (among patients with and without PCR testing),

*Pr(COVID-19*+, *Flu*+ | *Co-Tested)*: Probability that the individual received positive PCR tests for both COVID-19 and influenza within +/- 14 days of the index date (among patients with PCR tests available for both influenza and COVID-19).

Similarly, we define the following probabilities that PCR co-testing data is available:

*Pr(Co-Tested)*: Probability that the individual received PCR tests for both COVID-19 and influenza within +/- 14 days of the index date

*Pr(Co-Tested* | *COVID-19*+): Probability that the individual received PCR tests for both COVID-19 and influenza within +/- 14 days of the index date (among patients with a positive PCR test result for COVID-19),

*Pr(Co-Tested* | *Flu*+): Probability that the individual received PCR tests for both COVID-19 and influenza within +/- 14 days of the index date (among patients with a positive PCR test result for influenza),

*Pr(Co-Tested* | *COVID-19*+, *Flu*+): Probability that the individual received PCR tests for both COVID-19 and influenza within +/- 14 days of the index date (among patients with a positive PCR test results for both COVID-19 and influenza).

By definition,

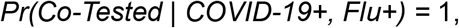

because all patients who have received positive PCR tests for both COVID-19 and influenza have received at least 1 PCR test of each type.

Let *n*_*co-tested*_ be the total number of individuals with PCR testing data available for both COVID-19 and influenza within +/- 14 days of the index date. Given these probabilities, we estimate the expected number of co-infection cases at the Mayo Clinic as:

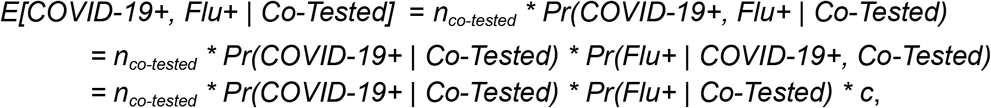

where:

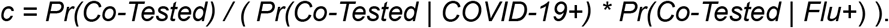

We can interpret this model parameter *c* as the ratio of co-testing PCR rates in the general population relative to the co-testing PCR rates in the COVID-19 and influenza positive populations. Substituting in the actual number of observed COVID-19 and influenza co-infection cases over the study period, we can solve for this model parameter *c* as:

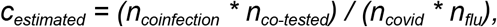

where:

*n*_*coinfection*_: number of co-infected cases,

*n*_*covid*_: number of cases with positive PCR tests for COVID-19, and

*n*_*flu*_: number of cases with positive PCR tests for influenza.

In order to estimate the expected number of co-infected cases for a particular time period *t* (e.g. from December 13, 2021 onwards), we used the same expected value formula as above with this estimated value for *c*:

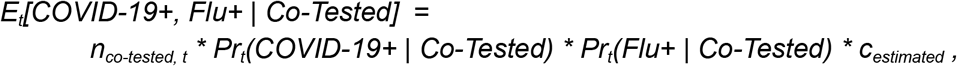

where: *n*_*co-tested, t*_, *Pr*_*t*_*(COVID-19*+ | *Co-Tested)*, and *Pr*_*t*_*(Flu*+ | *Co-Tested)* are time period-specific case counts and probability estimates.

### Statistical analysis

To identify clinical covariates enriched in the co-infected cohort with respect to the overall COVID-19 cohort, we report relative risk estimates for each of the categorical variables in the dataset, including: demographics, (sex, race, ethnicity), geographic location and time of PCR test, COVID-19 and influenza vaccination status, and comorbidities. For each covariate, the relative risk was computed as the rate in the co-infected cohort divided by the rate in the control cohort. In addition, we report 95% confidence intervals using the Delta method approximation^19^. Relative risks and 95% confidence intervals were computed using the “scipy” package (version 1.7.2) in Python.

## Results

### Recent surges in COVID-19 and influenza cases correspond with recent rises in the number of hospitalized patients with COVID-19 infection, influenza infection, and co-infections in the United States

In **Figure 1**, we show trends in COVID-19 and influenza infections, hospitalizations, and co-infection-related hospitalizations across the United States, aggregated from the CDC COVID Data Tracker, CDC FluView, and HHS Protect Public Data Hub. We observe that COVID-19 infection rates have risen to all time highs during the Omicron era (**Figure 1A**), while influenza infection rates have been negligible throughout the pandemic until recently at the beginning of the 2021-2022 flu season (**Figure 1B**). Although we are unable to observe co-infection rates in the overall US population, we can observe co-infection rates among hospitalized patients from HHS Protect data. In **Figures 1C** and **1D**, we show the average number of hospitalized patients with COVID-19 infection, influenza infection, and co-infections for 3,003 hospitals with data reporting to HHS Protect. We observe large peaks in hospitalized COVID-19 patients during the Original (Wuhan), Delta, and Omicron waves of the pandemic (**Figure 1C**). On the other hand, we observe that hospitalized influenza case counts were slightly elevated during the 2020-2021 flu season but have dramatically increased starting in October 2021 (**Figure 1D**). Case counts of hospitalized co-infected patients follow a similar trend, with a slight elevation during the 2020-2021 flu season and rising case counts observed from December 2021 to January 2022 (**Figure 1D**).

**Figure 1:**
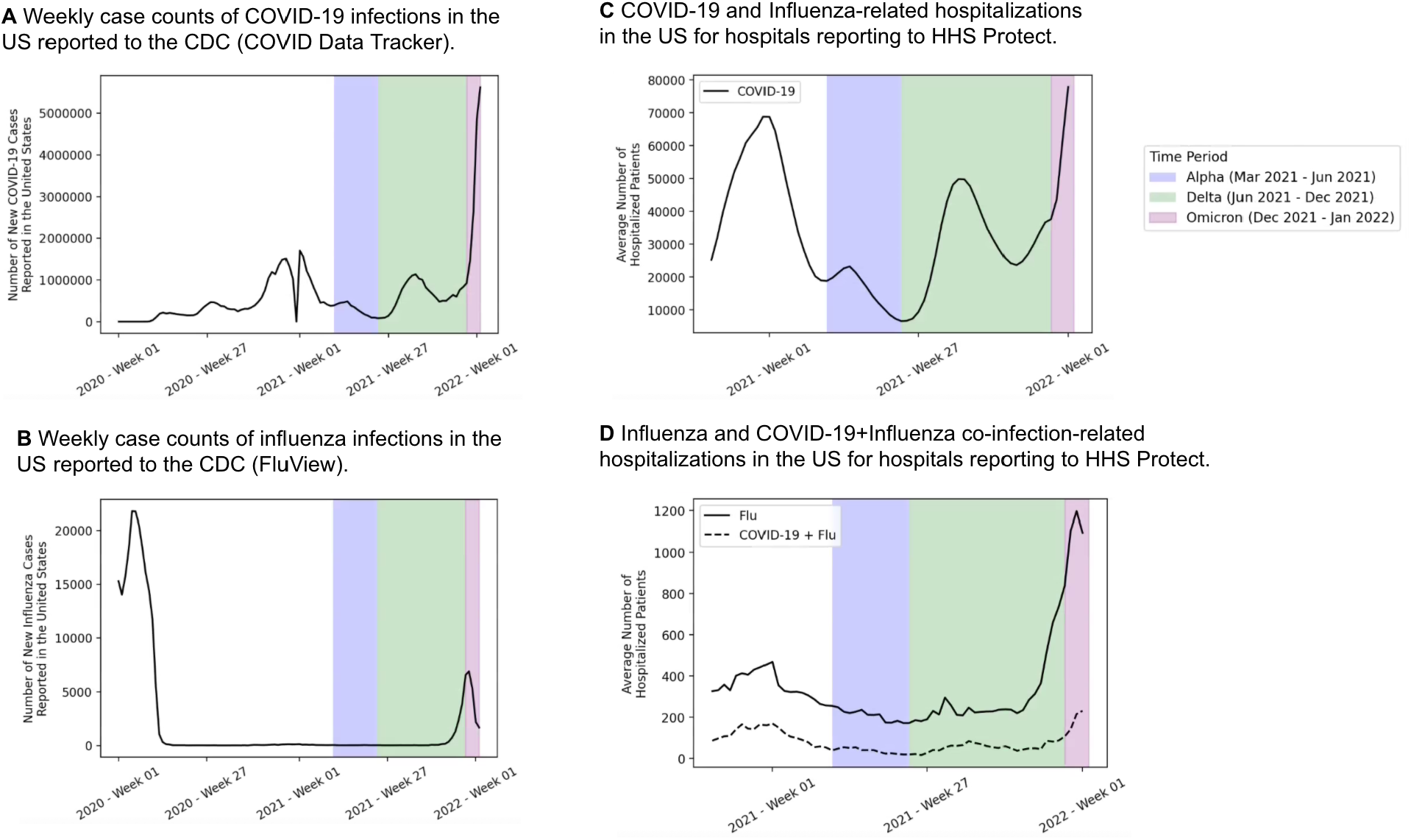
Case counts and hospitalizations for COVID-19, influenza, and co-infections in the United States. In panel (A), we show weekly case counts of new COVID-19 infections in the US reported by the CDC COVID Data Tracker. In panel (B), we show weekly case counts of new influenza infections in the US reported by CDC FluView. In panels (C) and (D), we show trends in hospitalized cases of COVID-19, Influenza, and co-infections over time for 3,003 hospitals in the U.S. which have submitted records to the HHS Protect Public Data Hub since the last week in October 2020 (see *Methods*). In panel (C), we show COVID-19 hospitalized case counts, and in panel (D) we show influenza and co-infection hospitalized case counts. In each of the plots, we shade the periods of time corresponding to the different waves of the pandemic, including Alpha (March 16, 2021 - June 15, 2021; **blue**), Delta (June 16, 2021 - December 13, 2021; **green**), and Omicron (December 14, 2021 - January 17, 2022; **purple**).

### The recent rise in hospitalized co-infection cases corresponds with the Omicron SARS-CoV-2 and H3N2 influenza strains

In the plots for **Figure 1**, we highlighted the time periods corresponding to different waves of the COVID-19 pandemic, including time periods when the Alpha, Delta, and Omicron variants were the most prevalent SARS-CoV-2 strains. Next, we provide a more granular view of the SARS-CoV-2 and influenza strains estimated to be in circulation in the United States over the course of the pandemic. In **Figure 2**, we provide the estimated percentages of individual SARS-CoV-2 and influenza strains sequenced in the United States based on the NCBI Virus and CDC FluVew databases, with SARS-CoV-2 strains including Alpha, Beta, Delta, Gamma, and Omicron, and influenza strains including H1N1 and H3N2. Taken in combination with the HHS Protect data, we observe that the recent rise in COVID-19 and influenza co-infection-related hospitalizations has occurred while the Omicron SARS-CoV-2 variant and the H3N2 influenza variant were most prevalent in the United States.

**Figure 2:**
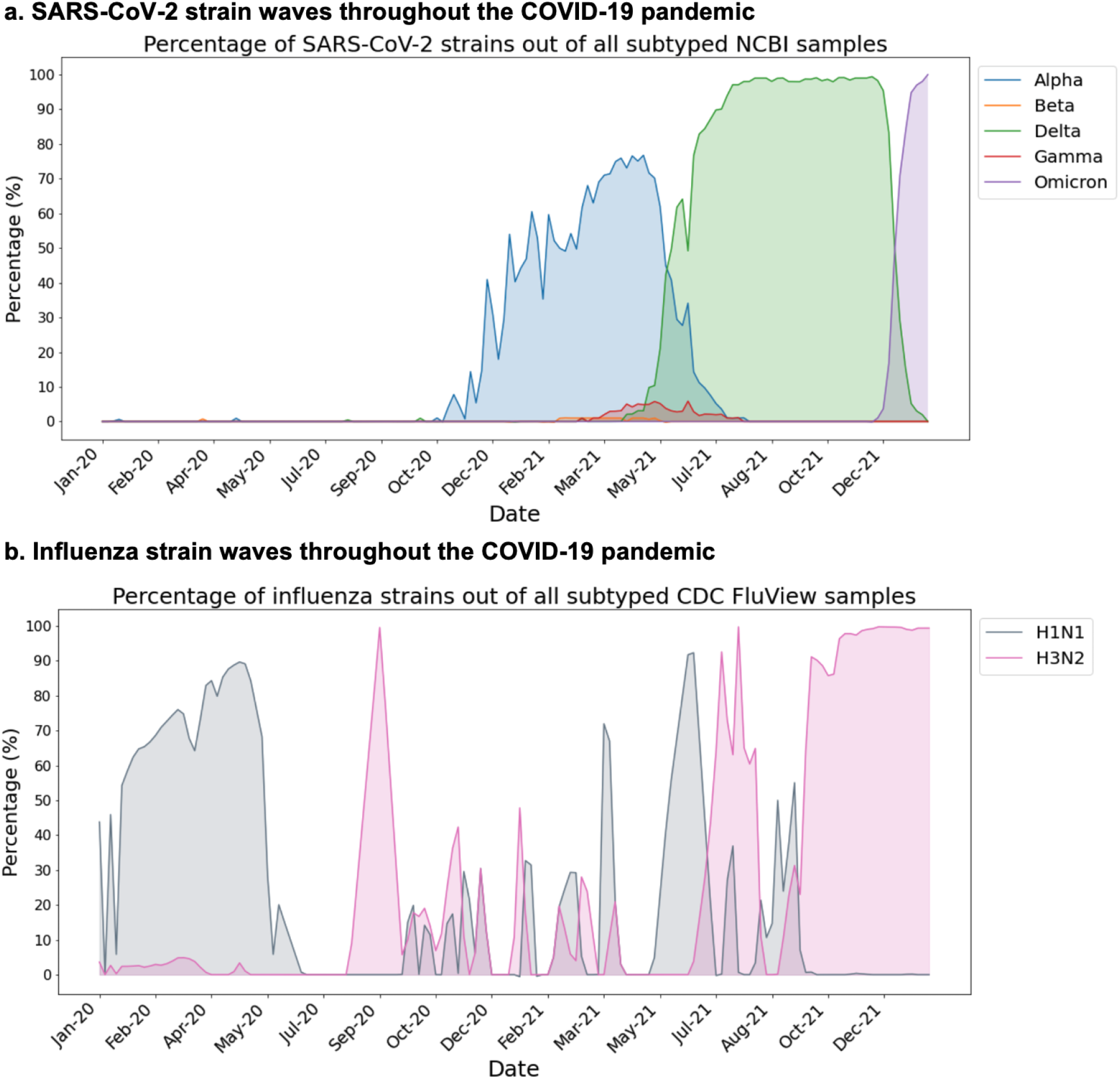
Trends of SARS-CoV-2 and influenza strain prevalences in the United States over time. In panel (A), we show SARS-CoV-2 strain percentages in the US from NCBI data, including the following strains: Alpha (**blue**), Beta (**orange**), Delta (**green**), Gamma (**red**), and Omicron (**purple**). In this plot, prevalence of the original (Wuhan) strain is not shown. In panel (B), we show influenza strain percentages in the US from CDC FluView data, including the following strains: A(H1N1)pdm09 (**gray**) and A(H3N2) (**pink**). In both panels (A) and (B), less common strains are omitted from the plots so the strain percentages do not necessarily add up to 100% each day.

### Hospitalized co-infection cases have been observed at low rates across the United States

In **Figure 3**, we show the geographic distribution of hospitalized COVID-19 and influenza cases in the United States on January 27, 2022 based on data from the HHS Protect Public Data Hub. We observe that the states with the highest numbers of hospitalized co-infection cases are Georgia (35 cases), Alabama (24 cases), and Texas (22 cases) (**Figure 3**). Alabama and Georgia have the highest percentage of hospitalized COVID-19 patients with influenza co-infections, but these percentages are still low (Alabama: 0.8% of hospitalized COVID-19 cases, Georgia: 0.7% of hospitalized COVID-19 cases). There are no states in which co-infections comprise over 1% of all COVID-19 hospitalized cases.

**Figure 3:**
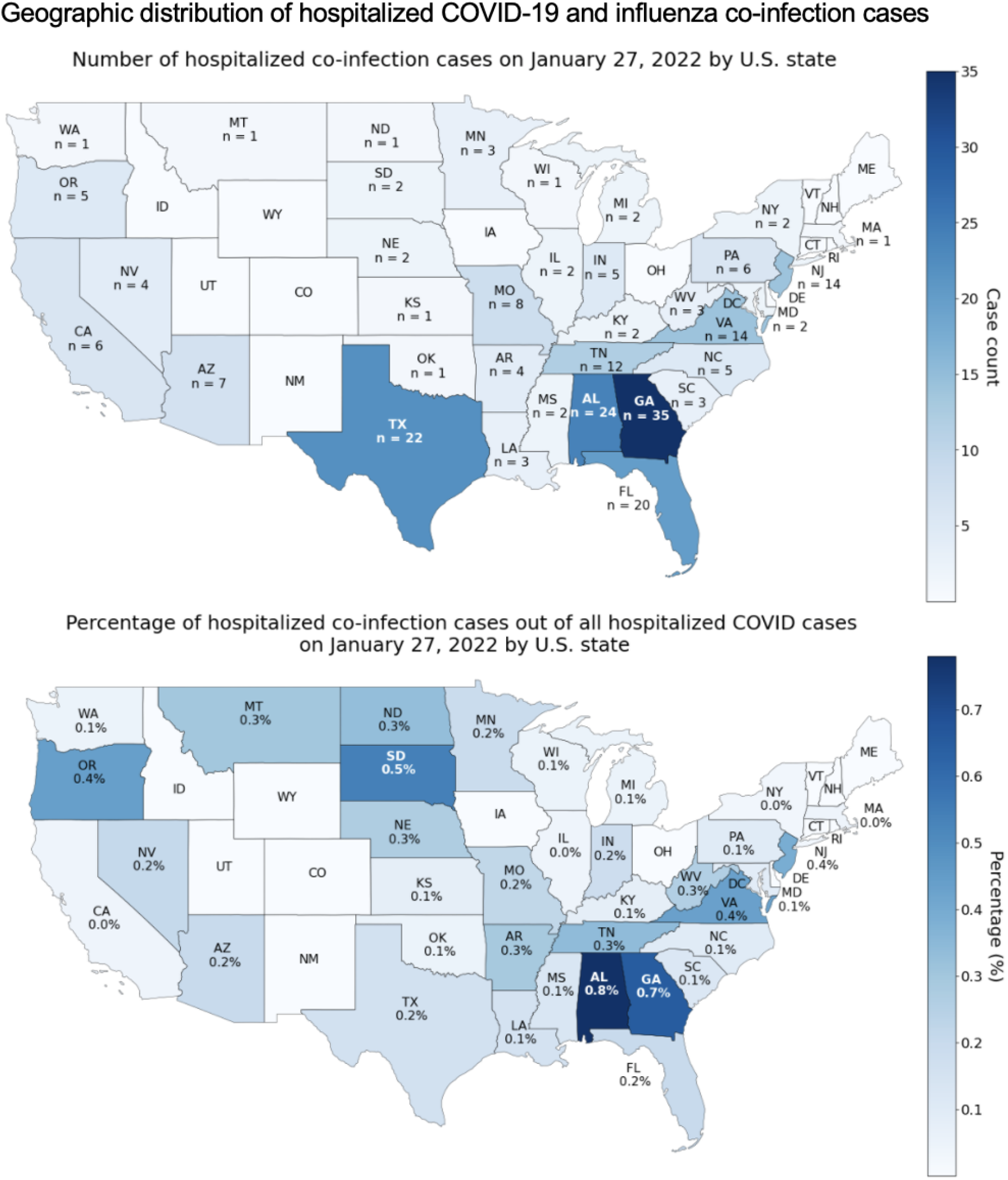
Geographic data on COVID-19 and influenza co-infections from HHS Protect Public Data Hub. We show the geographic distribution of hospitalized co-infection cases in the United States. The top map shows the states colored by total number of co-infection cases, and the bottom map shows the states colored by co-infection percentage. For each state, the co-infection percentage is calculated as the number of hospitalized co-infection cases divided by the total number of hospitalized COVID-19 cases.

### The recent surge in co-infection cases observed at the Mayo Clinic coincides with recent surges in both COVID-19 and influenza cases observed at the health system

EHR data from Mayo Clinic corroborates the observed trend in the public data that co-infection cases are rising in step with rises in COVID-19 and influenza (**Figure 4**). Co-infection cases were defined based on features in the EHR data as described in the methodology and summarized in **Figure 4A**. New cases of COVID-19 and influenza at Mayo Clinic (**Figures 4B** and **4C**) follow similar trends to those seen on the national level (**Figures 1A** and **1B**), and co-infection cases at Mayo Clinic coincide with the rise in the COVID-19 and influenza cases at the hospital system. In **Table S4**, we provide case counts of COVID-19 infections, influenza infections, and co-infections observed at the Mayo Clinic during each of the four study time periods (corresponding to the Wuhan (original), Alpha, Delta, and Omicron waves).

**Figure 4:**
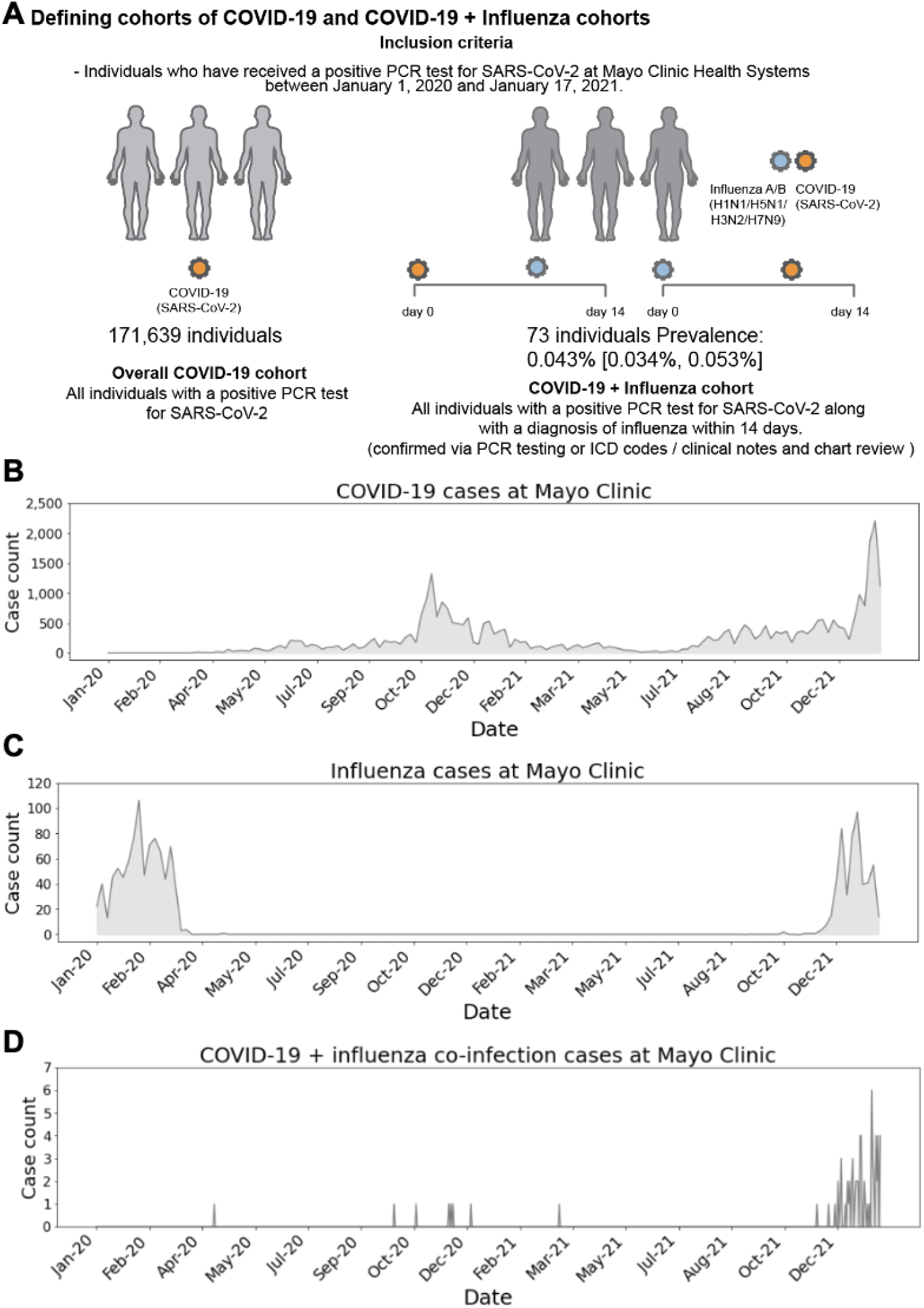
Schematic of cohort definitions and daily case counts of COVID-19 infection, influenza infection, and co-infections at the Mayo Clinic. In panel (A), we provide the inclusion criteria of COVID-19 and co-infection cohorts based on the Mayo Clinic EHR data and the resulting cohort sizes. In panels (B) and (C), we show counts of COVID-19 cases and influenza cases, respectively. In panel (D), we show counts of COVID-19 and influenza co-infection cases including 73 cases determined via PCR testing, ICD codes, and/or clinical notes. Note that the y-axis ranges in each plot are different, and the case counts for COVID-19 infection are significantly greater than the case counts for influenza infection which are in turn significantly greater than the case counts for COVID-19 and influenza co-infection.

### Laboratory testing for COVID-19 and influenza co-infections is low and confirmed co-infections are rare

Among 171,639 COVID-19 cases at the Mayo Clinic, only 10,431 (6.1%) cases had an influenza PCR test recorded within two weeks of their positive SARS-CoV-2 PCR test (**Table 1**). Of these 10,431 cases, only 63 individuals tested positive for influenza, resulting in an estimated co-infection prevalence of 0.604% (95% CI: [0.472%, 0.772%]). Given the possibility of incomplete lab data in the EHR, we also considered other documentation methods of influenza diagnosis, which increased the size of the co-infected cohort to 73 cases (3 additional cases determined from ICD codes, 7 additional cases determined from augmented curation of clinical notes). In the overall COVID-19 population (n = 171,639), the prevalence estimate of influenza co-infection from all three of these data sources is 0.043% (95% CI: [0.034%, 0.053%]) (**Table 1**). Among the population with PCR-confirmed influenza during the study period (n = 2,175), the prevalence of COVID-19 co-infection was 2.90% [2.27%, 3.69%] (**Table 1**).

**Table 1:** Case counts and estimated prevalences for COVID-19 and influenza co-infections from Mayo Clinic EHR data.

a) Case counts of co-infections from COVID-19 cohort.
| Cohort | Case count (%) |
| --- | --- |
| All COVID-19 cases (confirmed by PCR test) | 171,639 (100.0%) |
| COVID-19 cases with lab tests for influenza within +/- 14 days | 10,431 (6.1%) |
| COVID-19 and influenza co-infection within +/- 14 days, with diagnosis of influenza confirmed by:<br>- Lab test<br>- ICD-10 codes + Manual review<br>- Clinical notes + Manual review<br>- Any of the above | 63 (0.04%)<br>3 (0.00%)<br>7 (0.00%)<br>73 (0.04%) |

b) Case counts of co-infections from influenza cohort.
| Cohort | Case count (%) |
| --- | --- |
| All influenza cases during the study period<br>(January 1, 2020 to January 17, 2022) | 2,175 (100%) |
| COVID-19 and influenza co-infection within +/- 14 days, with diagnosis of influenza confirmed by:<br>- Lab test | 63 (2.9%) |

c) Prevalence estimates.
| Description | Cases / population | Prevalence (95% CI) |
| --- | --- | --- |
| Estimated prevalence of COVID-19 and influenza co-infections from overall COVID-19 cohort | 73 / 171,639 | 0.043% [0.034%, 0.053%] |
| Estimated prevalence of COVID-19 and influenza co-infections from COVID-19 cohort with lab tests available | 63 / 10,431 | 0.604% [0.472%, 0.772%] |
| Estimated prevalence of COVID-19 and influenza co-infections from overall Influenza cohort | 63 / 2,175 | 2.90% [2.27%, 3.69%] |

### Co-infection with SARS-CoV-2 and influenza is not more frequent than expected by chance

We next asked whether co-infections have occurred at unexpectedly high rates during various intervals throughout the pandemic, given the background rates of COVID-19 and influenza. For each time period analyzed, the observed co-infection case counts fall within or slightly below the 95% confidence intervals of the expected co-infection case counts (**Table 2**). In particular, during the Omicron era (from December 14, 2021 to January 17, 2022), the expected number of COVID-19 and influenza co-infection cases among this population was 80.9 cases (95% CI: [76.6, 85.1]), while the observed count was only 54 cases. During the Delta era (from June 16, 2021 to December 2021), the expected number of co-infection cases in this population was 13.9 (95% CI: [12.7, 15.2]), but only 9 co-infection cases were observed. These results suggest that after controlling for the background rates of both COVID-19 and influenza, the number of co-infections is not higher than expected by chance.

**Table 2:** Expected number of co-infections throughout the pandemic among patients at the Mayo Clinic with co-testing PCR data. In the first column, we show the total number of cases at the Mayo Clinic during the time period with PCR testing data available for both COVID-19 and influenza within +/- 14 days, including both positive and negative PCR tests. For each row, this total is used as the denominator to compute the percentages which are displayed next to case counts. In the middle columns, we show the number of cases with positive PCR tests for COVID-19, influenza, and both COVID-19 and influenza, respectively. In the last column, we show the expected number of cases with positive PCR tests for both COVID-19 and influenza, assuming that the probabilities of testing positive for COVID-19 and influenza are independent in the general population (see *Methods*). Note that for the first two time periods, all confirmed co-infection cases had influenza diagnoses determined via ICD codes or clinical notes and not lab tests, so the co-infection case counts for these time periods were zero.

| Time period | Cases with PCR testing for both COVID-19 and Flu within 14 days | COVID-19 positive tests<br>Case count (%) | Flu positive tests<br>Case count (%) | COVID-19 + Flu positive tests<br>Case count (%) | Expected number of COVID-19 + Flu positive tests<br>Case count (%)<br>[95% CI] |
| --- | --- | --- | --- | --- | --- |
| March 12, 2020 - March 15, 2021 | 14,243 | 1,167 (8.2%) | 155 (1.1%) | 0 (0.0%) | 2.5 (0.02%)<br>[2.1 (0.01%), 2.9 (0.02%)] |
| March 16, 2021 - June 15, 2021 | 1,806 | 42 (2.3%) | 0 (0.0%) | 0 (0.0%) | 0 (0.0%)<br>[0.0 (0.0%), 0.0 (0.0%)] |
| June 16, 2021 - December 13, 2021 | 28,771 | 3,909 (13.6%) | 516 (1.8%) | 9 (0.03%) | 13.9 (0.05%)<br>[12.7 (0.04%), 15.2 (0.05%)] |
| December 14, 2021 - January 17, 2022 | 25,447 | 6,752 (26.5%) | 1,535 (6.0%) | 54 (0.2%) | 80.9 (0.3%)<br>[76.6 (0.30%), 85.1 (0.33%)] |

### COVID-19 and influenza co-infections are associated with time and location of PCR test, re-infections, COVID-19 vaccine status, COVID-19 vaccine type, age, and sex

We next compared the clinical characteristics of the overall COVID-19 cohort (n = 171,639 cases) and the co-infected cohort (n = 73 cases) (**Table 3**). The co-infected cohort was younger on average, with a mean age of 28.4 years old (std dev: 21.7 years) compared to 39.8 years old (std dev: 20.9 years) for the overall COVID-19 cohort. In addition, co-infections were more prevalent among men compared to women (RR: 1.3, 95% CI: [1.1, 1.5]). Other clinical covariates such as race, ethnicity, and pre-existing conditions were similar between the two cohorts (**Table 3**). As expected, most co-infection cases (n = 55, 75.3%) occurred during the Omicron era (December 14, 2021 - January 17, 2022). Compared to the overall COVID-19 cohort, co-infection cases were more likely to be COVID-19 re-infections (RR: 2.2, 95% CI: [1.0, 6.0]) and to occur in individuals who were partially vaccinated (RR: 2.6, 95% CI: [1.3, 6.2]) or vaccinated with the Moderna COVID-19 vaccine (RR: 2.1, 95% CI: [1.2, 4.2]) (**Table 3**). We also observed that co-infection cases were slightly higher in the Mayo Clinic - Midwest region compared to the other regions (RR: 1.1, 95% CI: [1.0, 1.2]).

**Table 3:** Clinical characteristics of “COVID-19 + Flu” and “Overall COVID-19” cohorts at the Mayo Clinic. The “COVID-19 + Flu” cohort includes all individuals with a positive PCR test for SARS-CoV-2 and at least one of the following within 14 days: a positive laboratory test for influenza, an ICD code for influenza, or a clinical note indicating a diagnosis of influenza. The “Overall COVID-19” cohort includes all individuals with a positive PCR test for SARS-CoV-2.

| Clinical characteristic | COVID-19 + Flu cohort<br>Case count (%) | Overall COVID-19 cohort<br>Case count (%) | Relative risk<br>(95% CI) |
| --- | --- | --- | --- |
| Total number of cases | 73 | 171,639 |  |
| Type of SARS-CoV-2 infection<br>- Primary infection<br>- Re-infection | 69 (94.5%)<br>4 (5.5%) | 167,406 (97.5%)<br>4,233 (2.5%) | 0.97 [0.91, 1.02]<br>2.22 [1.01, 6.04]*** |
| Time of SARS-CoV-2 infection<br>- March 12, 2020 - March 15, 2021<br>- March 16, 2021 - June 15, 2021<br>- June 16, 2021 - December 13, 2021<br>- December 14, 2021 - January 17, 2022 | 7 (9.6%)<br>1 (1.4%)<br>10 (13.7%)<br>55 (75.3%) | 77,957 (45.4%)<br>7,673 (4.5%)<br>49,477 (28.8%)<br>36,532 (21.3%) | 0.21 [0.11, 0.44]***<br>0.31 [0.09, 2.21]<br>0.48 [0.28, 0.86]***<br>3.54 [3.09, 4.02]*** |
| COVID-19 vaccination status<br>- Unvaccinated<br>- Partial<br>- Full<br>- Boosted | 56 (76.7%)<br>5 (6.8%)<br>12 (16.4%)<br>0 (0.0%) | 137,880 (80.3%)<br>4,572 (2.7%)<br>24,093 (14.0%)<br>5,094 (3.0%) | 0.95 [0.84, 1.08]<br>2.57 [1.25, 6.24]***<br>1.17 [0.73, 1.99]<br>0.00 [0.00, 3.61] |
| Initial COVID-19 vaccine type<br>- None<br>- Janssen<br>- Moderna<br>- Pfizer/BioNTech | 56 (76.7%)<br>0 (0.0%)<br>8 (11.0%)<br>9 (12.3%) | 137,880 (80.3%)<br>2,272 (1.3%)<br>8,822 (5.1%)<br>22,665 (13.2%) | 0.95 [0.84, 1.08]<br>0.00 [0.00, 8.09]<br>2.13 [1.19, 4.21]***<br>0.93 [0.54, 1.76] |
| Flu vaccination status at time of SARS-CoV-2 infection<br>- Unvaccinated<br>- Vaccinated | 63 (86.3%)<br>10 (13.7%) | 153,795 (89.6%)<br>17,844 (10.4%) | 0.96 [0.87, 1.05]<br>1.32 [0.78, 2.39] |
| Site<br>- Mayo Clinic - Arizona<br>- Mayo Clinic - Florida<br>- Mayo Clinic - Midwest | 3 (4.1%)<br>4 (5.5%)<br>66 (90.4%) | 16,178 (9.4%)<br>17,570 (10.2%)<br>137,891 (80.3%) | 0.44 [0.18, 1.40]<br>0.54 [0.24, 1.45]<br>1.13 [1.04, 1.21]*** |
| Age at time of positive PCR test for SARS-CoV-2 (in years)<br>- Mean:<br>- Median:<br>- Std dev:<br>- IQR: | 28.4<br>21.7<br>21.7<br>(14.1, 41.2) | 39.8<br>38.3<br>20.9<br>(23.0, 56.0) |  |
| Sex<br>- Female<br>- Male<br>- Unknown / Non-binary | 28 (38.4%)<br>45 (61.6%)<br>0 (0.0%) | 87,472 (51.0%)<br>84,099 (49.0%)<br>68 (0.0%) | 0.75 [0.57, 1.01]<br>1.26 [1.05, 1.50]***<br>0.00 [0.00, 270.88] |
| Race<br>- Asian<br>- Black / African American | 2 (2.7%)<br>5 (6.8%) | 4,230 (2.5%)<br>7,295 (4.3%) | 1.11 [0.41, 4.64]<br>1.61 [0.78, 3.91] |
| - Native American<br>- Native Hawaiian / Pacific Islander<br>- White / Caucasian<br>- Other<br>- Unknown | 1 (1.4%)<br>0 (0.0%)<br>59 (80.8%)<br>5 (6.8%)<br>1 (1.4%) | 757 (0.4%)<br>287 (0.2%)<br>143,099 (83.4%)<br>6,911 (4.0%)<br>9,060 (5.3%) | 3.11 [0.94, 22.42]<br>0.00 [0.00, 64.04]<br>0.97 [0.86, 1.08]<br>1.70 [0.83, 4.13]<br>0.26 [0.08, 1.87] |
| Ethnicity<br>- Hispanic or Latino<br>- Not Hispanic or Latino<br>- Unknown | 7 (9.6%)<br>65 (89.0%)<br>1 (1.4%) | 12,773 (7.4%)<br>147,368 (85.9%)<br>11,498 (6.7%) | 1.29 [0.69, 2.68]<br>1.04 [0.95, 1.12]<br>0.20 [0.06, 1.48] |
| Elixhauser Comorbidity Index<br>- Mean:<br>- Std dev: | 0.7<br>5.4 | 1.7<br>6.5 |  |

## Discussion

Since the COVID-19 pandemic began in early 2020, cases of influenza have been dwarfed by cases of COVID-19. Several studies have shown that rates of influenza were lower in the 2020-2021 flu season, which has been largely attributed to social distancing measures for COVID-19^20–22^. In our study population with COVID-19, we found that the rate of lab testing for influenza co-infections was very low (6.1%). We expect that the low number of cases during the 2020-2021 flu season may explain why few influenza tests were ordered during the study time period, since most individuals would have a low pre-test probability for flu and a high pre-test probability for COVID-19. Starting in November 2021, the largest site started to systematically test COVID-19 on influenza swabs, but before this, testing was entirely dependent on provider suspicion. Even among individuals with both SARS-CoV-2 PCR testing and influenza PCR testing data available, we found that co-infection cases were rare, with an estimated prevalence of 0.604% (95% CI: [0.472%, 0.772%]) based upon a sample of 10,431 individuals. This is in line with previously estimated co-infection prevalences of 0.8% worldwide and 0.4% in the United States^3^.

Compared to the overall COVID-19 study population, the cohort of co-infected individuals at the Mayo Clinic had higher rates of several clinical covariates including: recent PCR diagnosis of SARS-CoV-2 (after December 14, 2021), geographic location (Mayo Clinic - Midwest), SARS-CoV-2 re-infection, COVID-19 vaccination status (partial), COVID-19 vaccine type (Moderna), younger age, and sex (male). Several of these variables such as time of PCR diagnosis and re-infection strongly depend upon time, and we observe that the case counts of co-infection at the Mayo Clinic were much higher in this past flu season compared to earlier in the pandemic. Each of these may be epi-phenomena of the timing of the flu season correlating with both the rise of Omicron and in an era of increasing reinfections in younger persons who were more likely to get vaccinated with the Moderna COVID-19 vaccine. Of note, these elevated co-infection case counts are in line with the expected numbers of co-infected cases given the background prevalences of COVID-19 and influenza in the Mayo Clinic population. From the epidemiological analysis of HHS Protect data, we observed that rates of co-infection-related hospitalizations have tracked closely with the rates of influenza-related hospitalizations in the United States. Together, this data suggests that recently observed increased rates of COVID-19 and influenza co-infections in the United States are most likely directly attributable to the recent surge in both COVID-19 and influenza cases during the 2021-2022 flu season rather than other factors such as the emergence of the Omicron variant. Aside from the time of SARS-CoV-2 infection, other factors may influence the risk of co-infection which could explain the enrichments among the co-infected cohort. For example, young males may be less likely to adhere to social distancing interventions, which would explain the higher prevalence of co-infections in this group. In addition, the higher rates of co-infections observed in the Mayo Clinic - Midwest region may be due to testing differences between the sites.

Although data sources from both HHS Protect and Mayo Clinic indicate that COVID-19 and influenza co-infection cases have surged in January 2022, there remains little data on disease severity for co-infected cases. Among the 23 co-infected cases observed at the Mayo Clinic with 30-day follow-up data available, there were no cases of hospitalization, ICU admission, or death observed (data not shown). This may be due to the co-infected population being younger (mean age: 34 years old) where severe disease is not common. Given that there is only a small population of co-infected cases with sufficient follow-up data available, it is difficult to draw conclusions on outcomes at this time. In the future, we plan to update this analysis to evaluate the clinical outcomes of the co-infected cohort in comparison to cohorts of patients with COVID-19 and influenza mono-infections. Going forward, it will be important to monitor the clinical outcomes of COVID-19 and influenza co-infection cases among a larger population of individuals with risk factors including older age, obesity, and immunocompromised status.

There are several important limitations to note for this study. First, this epidemiological analysis only includes data from US hospitals which have reported co-infections to the HHS Protect Public Data Hub. As a result, this dataset does not include all hospitals. Second, laboratory testing rates for influenza co-infections among COVID-19 cases are low, so the confidence interval for the prevalence of COVID-19 and influenza co-infections based on laboratory data alone is large. Third, in the probability model to estimate the expected number of co-infection cases at the Mayo Clinic, we assume that probabilities of COVID-19 infection and influenza infection are independent, but the true underlying probability distributions are most likely more complex. Indeed, given that the 2020-2021 flu season was extraordinarily mild due to the nonpharmaceutical interventions used to curb COVID-19, and compliance with these measures varied according to disease prevalence, there certainly is a complex interaction that cannot be fully modeled here. This is further complicated in the present flu season with re-implementation of nonpharmaceutical interventions in several localities during the present Omicron surge. Fourth, while we control for pre-existing conditions in the propensity matched analysis, currently we are not controlling for other factors such as respiratory symptoms at time of presentation which may impact testing rates for influenza and COVID-19 or the availability of a combined test. Finally, the EHR dataset only includes data on COVID-19 cases from a single healthcare system in the United States, which serves a patient population with a unique set of demographic and clinical characteristics. To assess prevalence and clinical outcomes for COVID-19 and influenza co-infections in the broader population, similar studies in other healthcare systems will be required.

Taken together, these data suggest that COVID-19 and influenza coinfection is occurring infrequently, and thus far co-infection rates during the Omicron era are no higher than expected given the background prevalences of both COVID-19 and influenza. Hospitalization with the combined illness appears to be infrequent, possibly attributed to the observed cases skewing younger than the overall COVID-19 population. Continued surveillance will be needed throughout the remainder of the influenza season.

## Data Availability

After publication, the data will be made available to others upon reasonable requests to the corresponding author. A proposal with detailed description of study objectives and statistical analysis plan will be needed for evaluation of the reasonability of requests. Deidentified data will be provided after approval from the corresponding author and Mayo Clinic.

## Supplementary Materials

**Table S1:** LOINC codes associated with influenza lab tests.

| LOINC code | Description | Influenza Type/Subtype |
| --- | --- | --- |
| 34487-9 | Influenza virus A RNA [Presence] in Specimen by NAA with probe detection | Influenza A |
| 40982-1 | Influenza virus B RNA [Presence] in Specimen by NAA with probe detection | Influenza B |
| 49521-8 | Influenza virus A H1 RNA [Presence] in Specimen by NAA with probe detection | Influenza A/H1N1 |
| 49524-2 | Influenza virus A H3 RNA [Presence] in Specimen by NAA with probe detection | Influenza A/H3N2 |
| 76077-7 | Influenza virus A RNA [Presence] in Bronchoalveolar lavage by NAA with probe detection | Influenza A |
| 76078-5 | Influenza virus A RNA [Presence] in Nasopharynx by NAA with probe detection | Influenza A |
| 76079-3 | Influenza virus B RNA [Presence] in Bronchoalveolar lavage by NAA with probe detection | Influenza B |
| 76080-1 | Influenza virus B RNA [Presence] in Nasopharynx by NAA with probe detection | Influenza B |
| 80382-5 | Influenza virus A Ag [Presence] in Upper respiratory specimen by Rapid immunoassay | Influenza A |
| 80383-3 | Influenza virus B Ag [Presence] in Upper respiratory specimen by Rapid immunoassay | Influenza B |
| 80588-7 | Influenza virus A M gene [Presence] in Nasopharynx by NAA with probe detection | Influenza A |
| 80589-5 | Influenza virus A H1 HA gene [Presence] in Nasopharynx by NAA with probe detection | Influenza A/H1N1 |
| 80590-3 | Influenza virus A H3 HA gene [Presence] in Nasopharynx by NAA with probe detection | Influenza A/H3N2 |
| 82166-0 | Influenza virus A RNA [Presence] in Nasopharynx by NAA with non-probe detection | Influenza A |
| 82170-2 | Influenza virus B RNA [Presence] in Nasopharynx by NAA with non-probe detection | Influenza B |
| 85477-8 | Influenza virus A RNA [Presence] in Upper respiratory specimen by NAA with probe detection | Influenza A |
| 85478-6 | Influenza virus B RNA [Presence] in Upper respiratory specimen by NAA with probe detection | Influenza B |
| 92141-1 | Influenza virus B RNA [Presence] in Respiratory specimen by NAA with probe detection | Influenza B |
| 92142-9 | Influenza virus A RNA [Presence] in Respiratory specimen by NAA with probe detection | Influenza A |
| 92976-0 | Influenza virus B RNA [Presence] in Lower respiratory specimen by NAA with non-probe detection | Influenza B |
| 92977-8 | Influenza virus A RNA [Presence] in Lower respiratory specimen by NAA with non-probe detection | Influenza A |

**Table S2:**
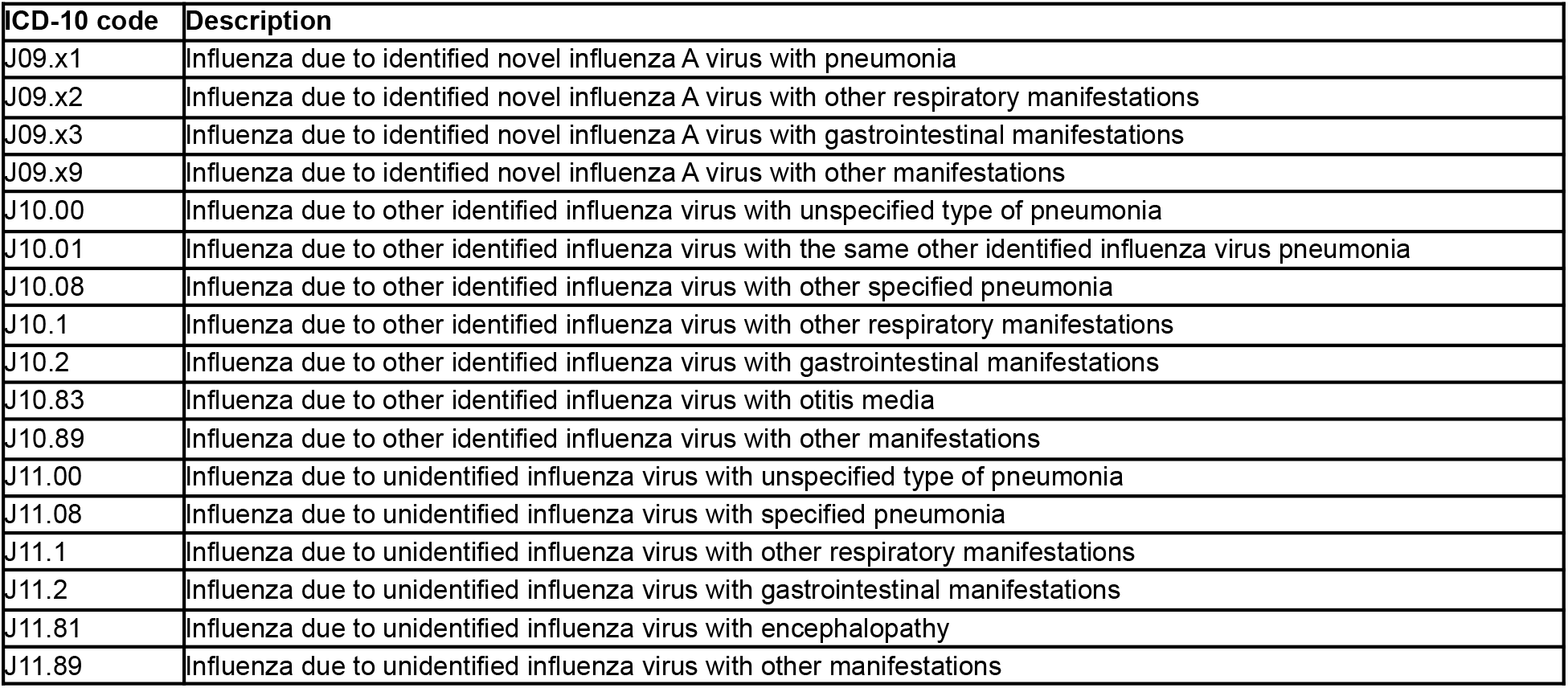
ICD-10 codes associated with influenza.

| ICD-10 code | Description |
| --- | --- |
| J09.x1 | Influenza due to identified novel influenza A virus with pneumonia |
| J09.x2 | Influenza due to identified novel influenza A virus with other respiratory manifestations |
| J09.x3 | Influenza due to identified novel influenza A virus with gastrointestinal manifestations |
| J09.x9 | Influenza due to identified novel influenza A virus with other manifestations |
| J10.00 | Influenza due to other identified influenza virus with unspecified type of pneumonia |
| J10.01 | Influenza due to other identified influenza virus with the same other identified influenza virus pneumonia |
| J10.08 | Influenza due to other identified influenza virus with other specified pneumonia |
| J10.1 | Influenza due to other identified influenza virus with other respiratory manifestations |
| J10.2 | Influenza due to other identified influenza virus with gastrointestinal manifestations |
| J10.83 | Influenza due to other identified influenza virus with otitis media |
| J10.89 | Influenza due to other identified influenza virus with other manifestations |
| J11.00 | Influenza due to unidentified influenza virus with unspecified type of pneumonia |
| J11.08 | Influenza due to unidentified influenza virus with specified pneumonia |
| J11.1 | Influenza due to unidentified influenza virus with other respiratory manifestations |
| J11.2 | Influenza due to unidentified influenza virus with gastrointestinal manifestations |
| J11.81 | Influenza due to unidentified influenza virus with encephalopathy |
| J11.89 | Influenza due to unidentified influenza virus with other manifestations |

**Table S3:** Codes associated with influenza vaccines.

| Vaccine code | Description |
| --- | --- |
| 146 | INFLUENZA VACCINE QUAD (FLUZONE/FLUARIX) (6 MONTHS AND OLDER)(PF) |
| 150 | IRB 17-005592 FLUZONE HIGH-DOSE OR STANDARD-DOSE INFLUENZA VACCINE |
| 155 | INFLUENZA VACCINE (FLUBLOK) (18 YEARS OR OLDER) (PF) |
| 161 | INFLUENZA VACCINE QUAD (FLUZONE) (6 MONTHS-35 MONTHS) (PF) |
| 185 | INFLUENZA VACCINE QV(FLUBLOK) (18 YEARS OR OLDER) (PF) |
| IIV | IRB- FLUARIX/FLUZONE INJ |
| IIV | flu vaccine qs 2015-16 (6mos+) Vial (PF) |
| IIV | flu vacc 2013-14 (36 mos+)(PF) Susp IM |
| IIV | flu vacc 2013-14 (6-35mos)(PF) Syringe I |
| IIV | flu vaccine 2012-13(3 yr+)(PF) Syringe I |
| IIV | flu vaccine qs 2014-15(6mos+) Susp IM |
| IIV | flu vacc 2013-14 (36 mos+)(PF) Syringe I |
| IIV | flu vacc ts 2013 (36 mos+)(PF) Susp IM |
| IIV | flu vac ts2013-14(36mo,up)(PF) Syringe I |
| IIV | flu vacc ts 2013 (6-35mos)(PF) Syringe I |
| IIV-HD | flu vacc ts 2014-15(65yr+)(PF) syringe I |
| IIV-HD | flu vacc ts 2013-14(65yr+)(PF) Syringe I |
| IIV-HD | flu vacc ts 2015-16(65yr+)(PF) syringe I |
| IIV3-HD | flu vacc ts2017-18 65yr up(PF) Syringe I |
| IIV3-HD | flu vacc ts2016-17 65yr up(PF) syringe I |
| IIV4 | flu vac qs 2015-2016 (PF) syringe IM |
| IIV4 | flu vaccine qs2016-17(6mos up) Susp IM |
| IIV4 | flu vaccine qs2017-18(6mos up) Susp IM |
| IIV4 | flu vac qs2016-17 36mos up(PF) Susp IM |
| IIV4 | flu vac qs2017-18 36mos up(PF) Syringe I |
| IIV4 | flu vac qs 2015-16(6-35mo)(PF) syringe I |
| IIV4 | flu vac qs2017-18 36mos up(PF) Susp IM |
| IIV4 | flu vac qs 2015-16(36mos+)(PF) Susp IM |
| IIV4 | flu vac qs 2016-17(6-35mo)(PF) syringe I |
| IIV4 | flu vac qs2016-17 36mos up(PF) syringe I |
| IIV4 | flu vac qs 2014-15(36mos+)(PF) syringe I |
| IIV4 | flu vac qs 2014-15(36mos+)(PF) Susp IM |
| IIV4 | flu vacc qs 2014 (6-35mos)(PF) syringe I |
| IIV4 | flu vac qs 2017-18(6-35mo)(PF) Syringe I |
| LAIV | flu vac qv live 2014 (2-49yrs) nasal spr |
| LAIV | flu vac 2013 (2-49yrs) Nasal Spray Syrin |
| LAIV | flu vac qv live 2015 (2-49yrs) nasal spr |
| null | Flublok (RIV3), 18-49yr |
| RIV3 | flu vac tv 2017(18yr up)rc(PF) Soln IM |
| RIV3 | flu vac tv 2014(18-49yr)rc(PF) Soln IM |
| RIV3 | flu vac tv 2015(18 yr+)rc(PF) Soln IM |
| RIV3 | flu vac tv 2013(18-49yr)rc(PF) Soln IM |
| RIV3 | flu vac tv 2016(18yr up)rc(PF) Soln IM |
| RIV4 | flu vac qv 2017(18yr up)rc(PF) Syringe I |

**Table S4:**
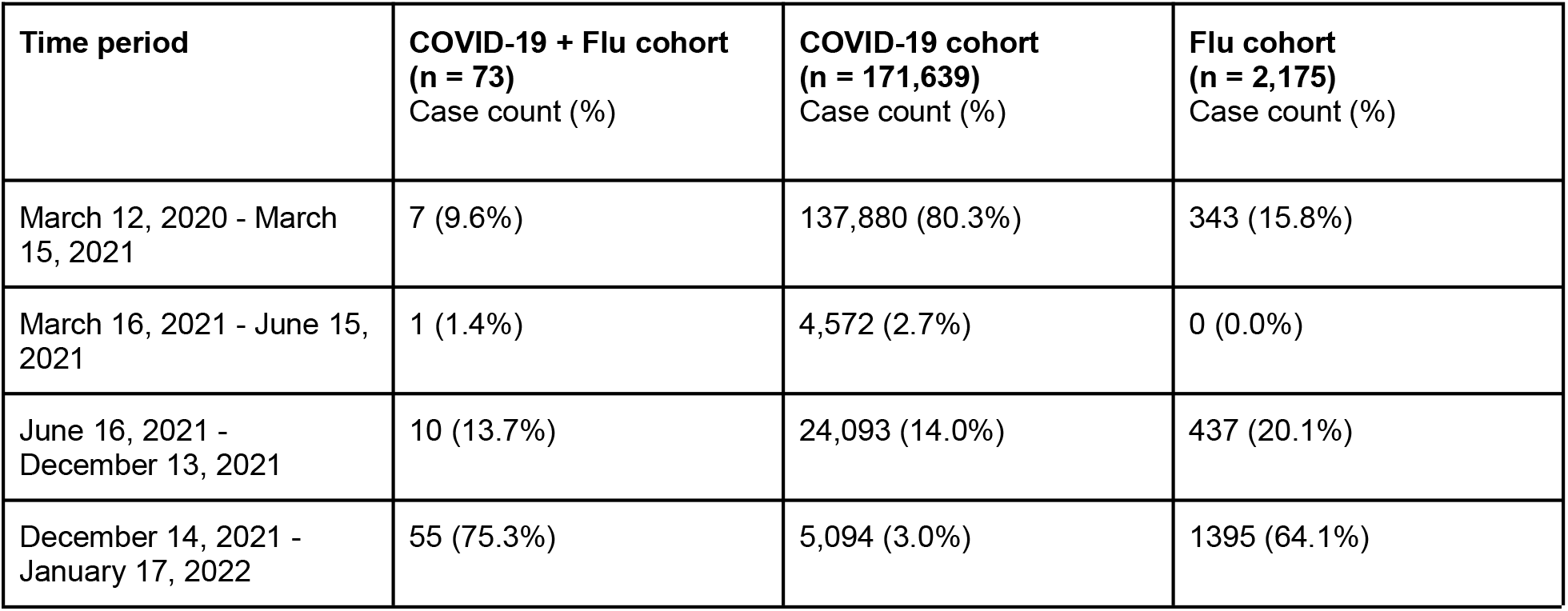
Prevalences of COVID-19 + influenza, COVID-19, and influenza throughout the pandemic based on Mayo Clinic EHR data. The “COVID-19 + Flu” cohort includes all individuals with a positive PCR test for SARS-CoV-2 and at least one of the following within 14 days: a positive PCR test for influenza, an ICD code for influenza, or a clinical note indicating a diagnosis of influenza. The “COVID-19” cohort includes all individuals with a positive PCR test for SARS-CoV-2. The “Flu” cohort includes all individuals with a positive PCR test for influenza.

| Time period | COVID-19 + Flu cohort<br>(n = 73)<br>Case count (%) | COVID-19 cohort<br>(n = 171,639)<br>Case count (%) | Flu cohort<br>(n = 2,175)<br>Case count (%) |
| --- | --- | --- | --- |
| March 12, 2020 - March 15, 2021 | 7 (9.6%) | 137,880 (80.3%) | 343 (15.8%) |
| March 16, 2021 - June 15, 2021 | 1 (1.4%) | 4,572 (2.7%) | 0 (0.0%) |
| June 16, 2021 - December 13, 2021 | 10 (13.7%) | 24,093 (14.0%) | 437 (20.1%) |
| December 14, 2021 - January 17, 2022 | 55 (75.3%) | 5,094 (3.0%) | 1395 (64.1%) |

